# Frontostriatal interactions and socioenvironmental associations with alcohol and cannabis onset in the Adolescent Brain Cognitive Development Study

**DOI:** 10.64898/2026.08.26.26360720

**Authors:** Karina A. Thiessen, Florence J. Breslin, Kara L. Kerr, Christian G. Schütz

**Affiliations:** Department of Psychiatry, Faculty of Medicine, University of British Columbia, Vancouver, BC, Canada; Djavad Mowafaghian Centre for Brain Health, Faculty of Medicine, University of British Columbia, Vancouver, BC, Canada; Rural Health, Oklahoma State University Center for Health Sciences, Tulsa, OK, USA; Hardesty Center for Clinical Research & Neuroscience, Oklahoma State University Center for Health Sciences, Tulsa, OK, USA; Department of Psychology, Oklahoma State University, Stillwater, OK, USA; British Columbia Mental Health and Substance Use Services, Provincial Health Services Authority, British Columbia, Canada

**Author notes:** **Corresponding author:** Karina A. Thiessen, 430 - 5950 University Blvd, Vancouver, BC, Canada, V6T 1Z3.

## Abstract

Adolescent substance use is a major public health concern due to increased risk of future physical and mental health conditions. Fronto-striatal functioning – particularly regarding inhibition and reward processing – may increase vulnerability to high-risk substance use. However, it remains unclear if these neurobiological differences precede substance use or are consequences of it. The ongoing Adolescent Brain Cognitive Development (ABCD) Study follows over 10000 youth, offering an unprecedented opportunity to longitudinally examine substance use patterns throughout development. We utilized family-clustered time-varying Cox proportional hazard models to prospectively examine main and interaction effects of right Inferior Frontal Gyrus (IFG) inhibitory control and bilateral nucleus accumbens (NAc) reward response, alongside early life adversity and peer substance use as predictors of alcohol and cannabis onset in the ABCD Study. We identified a significant crossover interaction such that left NAc activity had a slight positive association with first full alcoholic drink in the context of higher right IFG activity but a negative association in the context of lower right IFG activity. However, peer alcohol and cannabis use emerged as the strongest predictors of outcomes. Alcohol onset was also more common in females, and early life adversity was associated only with cannabis onset. Findings indicate that interactions between inhibition- and reward-related brain regions may impact risk for early substance use onset, but these effects may be modest relative to socioenvironmental factors. Additionally, divergent alcohol and cannabis findings suggest that risk profiles are substance specific. Peer-focused strategies should be considered in preventive efforts.

## 1. Introduction

Early adolescent substance use is linked to long-term negative outcomes, including future diagnosis of a substance use disorder[1,2], other psychiatric disorders and relational, academic, and employment difficulties[3]. For example, alcohol use before the age of 15 years old was previously found to increase the risk of an alcohol use disorder by 38% [4]. A twin study found that those who used cannabis by age 17 were up to five times more likely to use other substances or have a substance use disorder[5].

It is thought that these detrimental effects may be at least in part due to substance-induced disruption to neurodevelopmental changes in the adolescent brain. Use of addictive substances is known to alter fronto-striatal networks, largely due to their impact on dopaminergic systems in these networks [6]. Neurobiological sensitivity generally to substance effects is notably heightened during adolescence [7], and may be an explanatory mediating process by which adolescent substance use increases likelihood of a use disorder later in life. A recent analysis of the Adolescent Brain Cohort Study (ABCD) found that developmentally-normative improvements in cognitive functioning during adolescence were attenuated in youth that had previously used cannabis[8]. A 2026 systematic review found that age of first cannabis use was linked to subsequent differences in gray and white matter morphology, particularly in frontocortico-limbic regions[9]. Similar patterns have been linked to adolescent alcohol use [10].

However, while early substance use may have neurobiological consequences, neurobiological differences may also constitute predisposing vulnerabilities to adolescent substance use disorders. Early substance onset may be attributable to neurobiologically-based differences in inhibitory and reward-processing [7,11,12]. Specifically, anatomical and functional differences in the right inferior frontal gyrus (IFG) and bilateral nucleus accumbens (NAc), alongside corresponding differences in impulsivity and reward sensitivity, respectively, have been widely documented as increasing vulnerability to substance use-related problems. Reward sensitivity may contribute to increased salience or weighting of perceived substance-related rewards, alongside decreased salience or discounting of potential consequences. Deficits in cognitive control may impact ability to inhibit higher-risk behaviors, such as substance use, even in the face of negative consequences. Such studies identifying differences in frontostriatal circuitry associated with substance use-related difficulties have been fundamental to our current understanding of addiction [7,13,14].

It has also been proposed that interactions between brain regions primarily associated with “regulatory” executive functioning and brain regions primarily associated with reward/loss “reactive” processing (e.g. IFG and NAc interactions) should be considered when assessing neurobiological risk factors for substance-related problems [7,15]. From a neurodevelopmental perspective, imbalances in the developmental trajectories of frontocortical cognitive control and striatal reward sensitivity during adolescence further underscore the importance of examining interactions between these processes [7]. This may involve both top-down and bottomup processes. For example, if one is more responsive to perceived substance-related rewards, higher inhibitory control may be necessary to prevent substance use. Thus, cognitive control may impact susceptibility to substance use, particularly in those with heightened sensitivity to rewards. Such interactions may have greater impacts on substance use patterns compared to the role of each of these regions alone [15].

Lastly, it is generally agreed upon that both neurobiological and socioenvironmental factors likely play an etiological role in substance use disorders [16– 19]. Early life adversity has been well-characterized as a risk factor for early substance use [20,21]. Beyond adversity, peer substance use is also highly associated with adolescent substance use-related problems, independently of trauma exposure [22], and adolescent substance use behaviors tend to follow peer use patterns [23]. Yet, debates remain regarding the relative emphasis placed on the etiological role of biological versus socioenvironmental characteristics. The unique contributions of each have not been clearly delineated in parallel. While human experimental studies cannot be conducted to establish causality, large-scale prospective studies are needed to better temporally elucidate the distinct relationships between neurobiological and socioenvironmental factors and adolescent substance use [24,25].

Taken together, it remains unclear if differences in right IFG and NAc activity and interactions between these regions are either predisposing vulnerabilities or are consequences of adolescent substance use, and how neurobiological and socioenvironmental factors may uniquely contribute to risk. To address these gaps, we leverage the large-scale, longitudinal data from the Adolescent Brain Cognitive Development (ABCD) Study. The ABCD Study follows over 10,000 youth for 10 years, assessing a broad range of biopsychosocial characteristics, including psychopathology, substance use, familial characteristics, socio-environmental factors, genetics, and neurobiological factors. The study offers an unparalleled opportunity to rigorously investigate risk factors for adolescent substance use and identify potential targets for early intervention. Differences in gray matter morphology were previously found to predict early substance onset (primarily alcohol) up to age 14 in the ABCD study, but substance use and functional magnetic resonance imaging (fMRI) variables have not yet been investigated using the most recently-available data from the study.

Here, we prospectively examine individual associations and interactions between right IFG and bilateral NAc activity during response inhibition and reward anticipation as predictors of adolescent alcohol and cannabis use. In consideration of socioenvironmental factors we incorporate peer substance use and early life adversity in our analyses. Recent findings have also identified sex differences in substance use patterns in the ABCD study; thus, we also include sex as a covariate[26]. We hypothesized that each of these neurobiological and socioenvironmental factors would be independently associated with alcohol and cannabis use onset. We further predicted that right IFG activity would moderate the relationship between NAc activity and onset of alcohol and cannabis use.

### 2. Methods

We conducted a secondary analysis of tabulated data from the ongoing Adolescent Brain Cognitive Development (ABCD) Study (Data Release 7.0). Centralized institutional review board approval was obtained for the ABCD study from the University of California, San Diego. Written informed consent and assent was obtained from guardians and participants, respectively. Institutional review board approval for the present secondary analysis was obtained from the University of British Columbia in accordance with local requirements. We used all available data starting at baseline (age 9-10 years old) to year-7 follow-up (age 17-18 years old). Participants were excluded from corresponding alcohol or cannabis analyses if their age of first alcohol or cannabis use was less than or equal to their age at first study session (alcohol: 225; cannabis: 35). MRI data was collected biennially starting at baseline and all other data was collected annually.

### 2.1. Functional Magnetic Resonance Imaging (fMRI)

Participants completed fMRI scans biennially, starting at baseline. We utilize tabulated biennial data from the Monetary Incentive Delay (MID) task, the Stop Signal Task (SST). Data that did not meet ABCD study quality control recommendations were excluded [27]. We describe the tasks briefly here but more information on MRI task, sequence, processing, region of interest parcellation, and quality control methods have been previously well documented and can be found in the ABCD Study Documentation [27], Casey et al. [28], and Hagler et al. [29].

#### 2.1.1. Monetary Incentive Delay (MID) Task

The MID task assesses reward processing [30,31]. The task comprises trials involving the presentation of a large or small reward or loss, or neutral cue; followed by a target cue, during which participants must respond to a target cue to win the reward or avoid a loss; and then response feedback. The MID task has been found to elicit a response in the ventral striatum/NAc during reward processing and to be linked to substance addiction [28,30,32–34]. We examined left and right NAc anticipation of reward versus neutral contrasts from the task.

#### 2.1.2. Stop Signal Task (SST)

The SST assesses response inhibition, defined as the ability to cancel or stop an already-initiated action, and is associated with activity in the right IFG and with addiction [28,35–37]. The SST consists of “go” trials and “stop” trials. During “Go” trials, response cues (left or right-facing arrows) are presented for less than 1000ms, during which participants must press a button corresponding to the direction of the arrow. During “stop” trials, response cues are presented for less than 900ms, followed by a “stop” signal (upward facing arrow) for approximately 300ms. Participants must inhibit their response when a stop signal is presented. The stop signal delay is computed as the time between the onset of the initial response cue and the stop signal. The stop signal delay is continuously adjusted according to task performance, such that the delay is reduced by 50ms after an unsuccessful inhibition or increased by 50ms after a successful inhibition. We used right pars opercularis IFG [38] correct stop versus correct go contrasts from the task.

### 2.2. Early Life Adversity

Annual composite ELA scores were computed according to the “ELA+” methods in Breslin et al. [39]. Code was updated according to ABCD Data Release 7.0. Domains incorporated in the composite scores include physical, emotional, and sexual abuse; physical and emotional neglect; at-home violence; caregiver divorce; caregiver psychiatric illness; caregiver substance misuse; caregiver incarceration; community disaster and violence exposure; familial financial difficulties; and caregiver separation. Sources included caregiver and youth reported measures. As age and sex-normalized T-scores for caregiver psychiatric assessments were not available, within-sample ABCD caregiver T-scores were computed and used in composite ELA score calculations.

### 2.3. Peer Substance Use

Participant-reported peer alcohol and cannabis use data were collected annually [40–43]. Peer alcohol and cannabis use were each scored as a binary (none or a few/some/most/all) according to youth responses to the questions “How many of your friends drink alcohol (full beer, wine, or liquor?)” and “How many of your friends use cannabis?”, respectively.

### 2.4. Substance Use

Alcohol and cannabis use data were collected biannually via youth self-report. Time to first alcohol and cannabis use were extracted from summary substance use data provided in the study data release and was defined by the age at which the participant first used any form of alcohol or cannabis containing product. We examined time to first full alcoholic drink or time to first cannabis use for initiation (excludes low level use such as a “sip” or “puff”). Initiation was coded as a binary (yes/no) based on provided summary data. See Lisdahl et al. [42] for additional details on substance use data collection methods.

### 2.5. Cox Regression Analyses

Family-clustered time-varying robust Cox proportional hazard models were computed to evaluate the relationships between fMRI measures and each of alcohol and cannabis initiation. We tested interactions between (1) left NAc MID and right IFG SST activity and (2) right NAc MID and right IFG SST activity. Main effects were also tested in each model. Preliminary correlations between right IFG activity and NAc activity were non-significant, verifying that these capture distinct constructs (*p* > .05). ELA composite scores, peer substance use and fMRI variables were included as timevarying predictors. Sex assigned at birth was included as a time-invariant predictor. Peer alcohol and cannabis use, assessed via the questions “How many of your friends drink alcohol/use cannabis?” were converted into binary variables (none or a few/some/most/all) at each timepoint. Continuous predictors were standardized into zscores calculated by timepoint. Models were clustered by family to account for nonindependence of siblings. For outcome variables, stop time was input into the models as age of first initiation, or most recent follow-up year in which data was collected in the case of censored outcomes (i.e., no reported initiation). We prioritize effect sizes and confidence intervals, beyond nominally reported *p*-values, in interpretation of findings given the large sample size. All analyses and figures were generated in RStudio [44– 48].

For those reporting alcohol or cannabis initiation, only data from predictors from timepoints *preceding* their corresponding substance use event were included. Participants for whom their age of first use was before their age at their first included observation were excluded from analyses. All available data from each timepoint was otherwise used. Last observation carried forward (LOCF) was used in the case of missing data or fMRI scans that did not meet ABCD study quality control recommendations [27]. The assumption of proportional hazards was violated when proportional hazards tests on Schoenfeld residuals were conducted (*p*’s < .05) and confirmed via visual inspection; however, the violation was deemed negligible upon finding no meaningful differences in results when splitting variables by time. Prior literature has found that effects of such violations are minimal [49].

## 3. Results

### 3.1. Demographics and Descriptives

Our analyses comprised *n =* 10148 and 10338 for alcohol and cannabis, respectively, out of the original ABCD Study sample of *N =* 11875. Mean age of alcohol and cannabis initiation were 14.80 (1.62) and 14.88 (SD = 1.41), respectively. Further descriptives are detailed in Table 1.

**Table 1.**
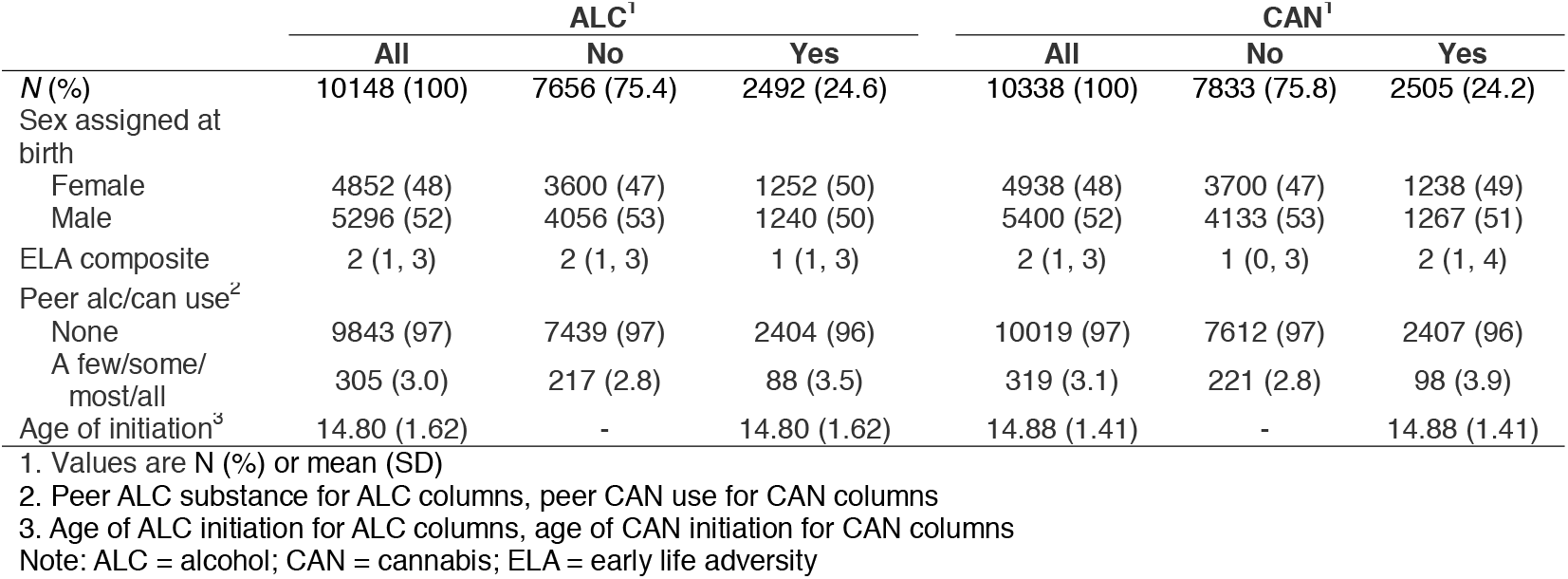
Descriptives.

### 3.2. Alcohol and Cannabis Initiation

Findings for alcohol and cannabis initiation, including statistics, are shown in Figure 1. Findings for covariates were consistent between right IFG-left NAc and right IFG-right NAc alcohol and cannabis models. Scanner site was explored as an additional covariate but did not meaningfully change findings and was thus removed from our models.

**Figure 1.**
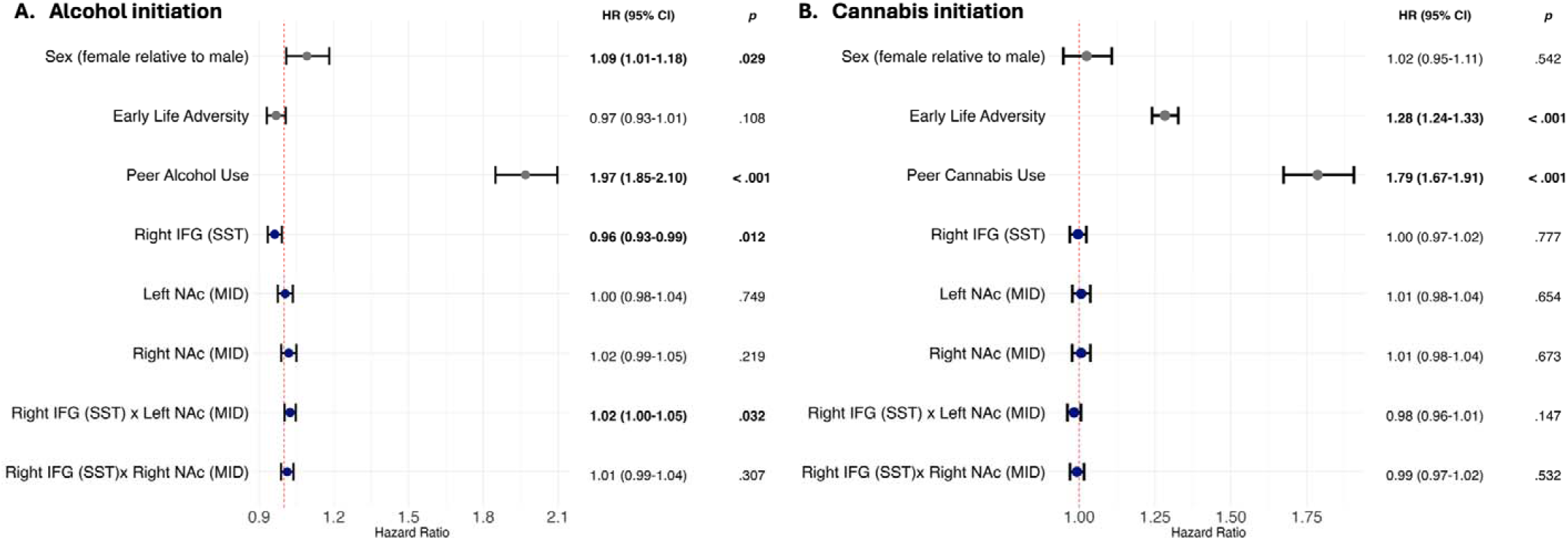
Cox proportional hazard model results. **(A)** Alcohol initiation. **(B)** Cannabis initiation. Covariate findings were consistent across right IFG-left NAc and right IFG-right NAC models; thus, hazard ratios from one model are shown in gray to avoid redundancy. Hazard ratios for neuroimaging measures are indicated in dark blue. CI = Confidence interval; HR = hazard ratio; IFG = inferior frontal gyrus; NAc = nucleus accumbens; MID = Monetary Incentive Delay Task; SST = Stop Signal Task.

Female participants had higher hazards of alcohol initiation. Peer alcohol use increased hazards of alcohol initiation and had the strongest effect in our models. ELA scores did not significantly predict alcohol initiation. There was a significant main effect of right IFG activity and a significant crossover interaction between right IFG and left NAc activity. Post-hoc analyses revealed that left NAc activity displayed a negative trend with initiation in the context of lower right IFG activity but inverted to a positive trend in the context of higher right IFG activity (Figure 2).

**Figure 2.**
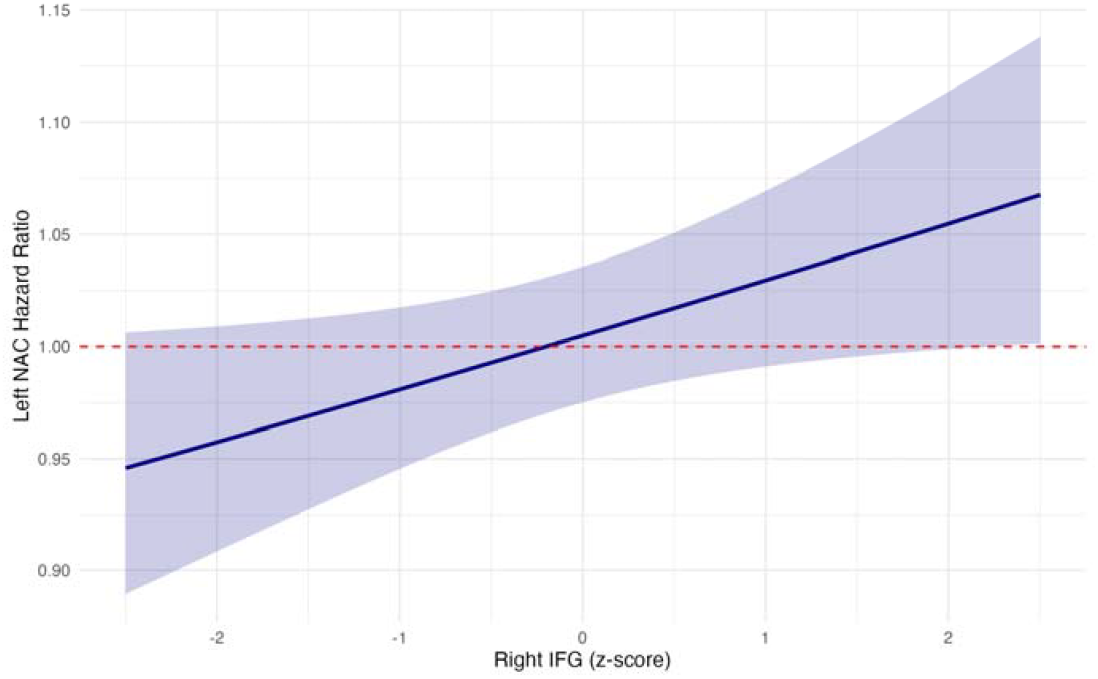
Hazard ratios of alcohol initiation for left NAc (MID) by right IFG (SST) activity. Shading indicates 95% confidence intervals. IFG = inferior frontal gyrus; NAc = nucleus accumbens; SD = standard deviation.

There were no significant sex differences for cannabis initiation. Similarly to our alcohol models, peer cannabis use increased hazards of cannabis initiation, and had the strongest association to cannabis initiation. ELA scores were associated with higher hazards of cannabis initiation. There were no significant relationships with NAc or right IFG activity.

## Discussion

In this study, we address the need for prospective investigations on risk factors for adolescent substance use, capitalizing on the ABCD Study’s large-scale, longitudinal data obtained from a diverse national sample. We explored both neurobiological and socioenvironmental risk factors in parallel, allowing for the delineation of the potential unique contributions of these different factors. By using time-varying analysis, we were able to evaluate variables of interest dynamically, leveraging all available datapoints across timepoints while still maintaining a prospective approach. Family-clustering enabled the inclusion of siblings, accounting for potential collinearity between observations and allowing us to retain a larger proportion of the original sample.

Prevalence of alcohol and cannabis initiation were high, at approximately 24%. Results often differed between alcohol and cannabis outcomes, which may indicate etiological heterogeneity in adolescent substance use patterns. Right IFG and NAc activity were unrelated to alcohol and cannabis initiation. There was a significant interaction between right IFG inhibitory control-related and left NAc reward anticipation-related activity in predicting alcohol initiation, providing some support for the neurocognitive regulation-reaction hypothesis [15]. We found that left NAc activity – generally linked to increased reward sensitivity – was positively associated with alcohol initiation in the context of increased right IFG activity. Heightened right IFG activity during response inhibition may be indicative of a compensatory response and heightened effortfulness of cognitive control (i.e., lower inhibitory executive functioning) [15,50]. Brain activations during the SST have been previously found to be negatively correlated with behavioral task performance [51]. In contrast, left NAc activity in the context of reduced right IFG activity was negatively associated with alcohol initiation, suggesting that increased cognitive control may be protective against alcohol use in those with heightened NAc reward sensitivity. This potential moderating relationship also indicates that substance use onset may involve both bottom-up (“reactive”) and top-down (“regulatory”) processes. However, while the interaction between right IFG and NAc activity in relation to initiation was statistically significant, this should be interpreted cautiously with consideration for confidence intervals.

Beyond this interaction in alcohol initiation, neuroimaging-based measures were largely unrelated to substance use outcomes. As substance use was examined prospectively in our analyses, our findings suggest that previously identified differences in NAc and right IFG function found in the context of substance use may arise later in development, emerge as a consequence of escalating substance use, and/or only predict higher threshold substance use, as opposed to age of first initiation. It is possible that other regions of interest or other neuroimaging approaches not investigated in the present analyses may play a role in risk for adolescent substance use. Further research investigating main and interaction effects in relevant brain regions may be warranted, especially given the lower-threshold substance use metric used in our analyses. However, a prior study of neurobiological and contextual risk factors of alcohol initiation utilizing earlier waves of data from the ABCD Study found that statistical models including gray matter morphology in the NAc and prefrontal cortex did not meaningfully predict alcohol initiation when accounting for contextual factors. Taken together, the relative impact of these neurobiological processes may be less meaningful in comparison to other risk factors for alcohol and cannabis onset by age 18.

Female participants were more likely to engage in alcohol initiation than male participants. This sex difference did not replicate when examining cannabis outcomes. Historically, males have been more likely to drink alcohol [52] and use cannabis [53,54]. However, our finding of increased risk of alcohol use for female participants parallels with year-6 follow-up reports from the ABCD study [26] and recent trends in adolescent alcohol use in the United States [55]. The lack of a significant sex difference in cannabis outcomes may reflect the increasing prevalence of use amongst females [53]. While the ABCD study is in the United States, findings also converge with trends in sex-differences in alcohol and cannabis use in Canada [56,57]. While the long-term outcomes of demographic shifts in substance use are unknown, this may have implications for potential changes in substance use disorder demographics in the future[26].

ELA was positively associated with of cannabis use, but not alcohol use. Interestingly, while non-significant, the relationship between ELA and alcohol initiation was trending in the opposite direction compared to cannabis initiation, further underscoring that early alcohol and cannabis use may be driven by different processes. A prior study found that while ELAs were associated with both alcohol and cannabis use, internalizing and externalizing problems mediated the relationship between ELA and cannabis use, but not alcohol use [20]. ELAs were also found to be more strongly related to adolescent cannabis use frequency than alcohol use frequency. Previous studies have also identified positive associations between SES and alcohol outcomes [58–61], but negative associations with other substances [60]. It is also important to note that lower SES may be related to more substance-use related harms, regardless of consumption levels [62]. Further research is needed to examine the nuances between adversity and substance use. Psychosocial interventions aimed at directly preventing or minimizing impacts of adversity should also be considered. While our evidence indicates that these would be particularly relevant to early cannabis use, and less-so for alcohol use, ELAs are broadly associated with adverse health and psychosocial outcomes, and interventions may therefore have widespread benefits.

Peer alcohol and cannabis use were the strongest predictors of age of onset, increasing risk by 47-97%. This relationship has been frequently identified, highlighting the often social nature of early substance use [63–67]. This relationship may be mediated by increased perceived positive expectancies around substance effects [68]. In consideration of the interaction between right IFG-left NAc in our findings, perceived positive expectancies may be particularly relevant to substance use risk for those that have stronger left NAc responses to reward and right IFG responses during inhibitory processing; although, this would need to be tested empirically. Peer- and communitybased strategies may be an effective approach to adolescent substance use prevention and intervention [67,69,70]. Also, while we identified substance use behaviors, prospectively (i.e., *after* peer substance use), there is prior evidence that this may be a bidirectional relationship [63,66], and peer-based interventions may thus have broader reach.

This study has some limitations. While we focused on the right IFG and NAc, other regions, such as the amygdala, insula, and fronto-cortical regions, likely warrant investigation [28,71,72]. Response to rewards and losses, or other neurocognitive functions, could also be considered. Secondly, while we examined substance use prospectively, etiology cannot be conclusively determined in an observational study. Lastly, the ABCD Study is subject to sampling biases, and may not be generalizable to other populations [73,74].

In summary, alcohol and cannabis initiation have somewhat distinct risk profiles, with differing socioenvironmental and neurobiological risk factors. Peer substance use was the most consistent and strongest predictor across both substances. Top-down and bottom-up interactions between brain regions associated with cognitive control and reward processing may be relevant in the context of alcohol initiation, but interpretability is limited by relatively broad confidence intervals. Peer-based strategies and psychosocial approaches to reducing early life adversity may be particularly important in preventing higher-risk adolescent substance use. Future directions include investigating other neuroimaging-based factors or higher threshold substance use. To our knowledge, this is the largest study to prospectively examine socioenvironmental and neurobiological correlates of substance use onset in adolescence using the most current data from the ABCD Study.

## Data Availability

Data from the Adolescent Brain Cognitive Development (ABCD) Study can be requested via the NIH Brain Development Cohorts Data Sharing Platform (https://www.nbdc-datahub.org).

https://doi.org/10.82525/8f3w-5260

https://www.nbdc-datahub.org

https://abcdstudy.org

## 5. Acknowledgments

Data used in the preparation of this article were obtained from the Adolescent Brain Cognitive Development− (ABCD) Study, held in the NIH Brain Development Cohorts Data Sharing Platform. This is a multisite, longitudinal study designed to recruit more than 10,000 children aged 9–10 and follow them over 10 years into early adulthood. The ABCD Study® is supported by the **National Institutes of Health** and additional federal partners under award numbers:

U01DA041048, U01DA050989, U01DA051016, U01DA041022, U01DA051018, U01D A051037, U01DA050987, U01DA041174, U01DA041106, U01DA041117, U01DA0410 28, U01DA041134, U01DA050988, U01DA051039, U01DA041156, U01DA041025, U0

1DA041120, U01DA051038, U01DA041148, U01DA041093, U01DA041089, U24DA04 1123, U24DA041147. A full list of supporters is available at Federal Partners – ABCD Study.

ABCD consortium investigators designed and implemented the study and/or provided data but did not necessarily participate in the analysis or writing of this report. This report reflects the views of the authors and may not reflect the opinions or views of the NIH or ABCD consortium investigators. The ABCD data repository grows and changes over time. The ABCD data used in this report came from doi: https://doi.org/10.82525/8f3w-5260. DOIs can be found at https://nda.nih.gov/abcd/abcd-annual-releases.

## References

[1] Behrendt S, Wittchen H-U, Höfler M, Lieb R, Beesdo K. Transitions from first substance use to substance use disorders in adolescence: Is early onset associated with a rapid escalation? Drug Alcohol Depend 2009;99:68–78. 10.1016/j.drugalcdep.2008.06.014.

[2] Leung J, Chan GCK, Hides L, Hall WD. What is the prevalence and risk of cannabis use disorders among people who use cannabis? a systematic review and meta-analysis. Addict Behav 2020;109:106479. 10.1016/j.addbeh.2020.106479.

[3] Poudel A, Gautam S. Age of onset of substance use and psychosocial problems among individuals with substance use disorders. BMC Psychiatry 2017;17:10. 10.1186/s12888-016-1191-0.

[4] Dawson DA, Goldstein RB, Patricia Chou S, June Ruan W, Grant BF. Age at First Drink and the First Incidence of Adult-Onset DSM-IV Alcohol Use Disorders. Alcohol Clin Exp Res 2008;32:2149–60. 10.1111/j.1530-0277.2008.00806.x.

[5] Lynskey MT, Heath AC, Bucholz KK, Slutske WS, Madden PAF, Nelson EC, et al. Escalation of Drug Use in Early-Onset Cannabis Users vs Co-twin Controls. JAMA 2003;289:427–33. 10.1001/jama.289.4.427.

[6] Volkow ND, Blanco C. Substance use disorders: a comprehensive update of classification, epidemiology, neurobiology, clinical aspects, treatment and prevention. World Psychiatry 2023;22:203–29. 10.1002/wps.21073.

[7] Casey BJ, Jones RM. Neurobiology of the Adolescent Brain and Behavior: Implications for Substance Use Disorders. J Am Acad Child Adolesc Psychiatry 2010;49:1189–201. 10.1016/j.jaac.2010.08.017.

[8] Wade NE, Sullivan RM, Wallace AL, Visontay R, Szpak V, Lisdahl KM, et al. Longitudinal neurocognitive trajectories in a large cohort of youth who use cannabis: combining self-report and toxicology. Neuropsychopharmacology 2026;51:1546–55. 10.1038/s41386-026-02395-1.

[9] Ricci V, Sarni A, De Berardis D, Martinotti G, Maina G. How adolescent cannabis use reshapes the developing brain — a systematic review. Front Psychiatry 2026;17. 10.3389/fpsyt.2026.1822300.

[10] Lees B, Meredith LR, Kirkland AE, Bryant BE, Squeglia LM. Effect of alcohol use on the adolescent brain and behavior. Pharmacol Biochem Behav 2020;192:172906. 10.1016/j.pbb.2020.172906.

[11] Green R, Meredith, Lindsay R., Mewton L, Squeglia LM. Adolescent Neurodevelopment Within the Context of Impulsivity and Substance Use. Curr Addict Rep 2023;10:166–77.

[12] Hamidullah S, Thorpe HHA, Frie JA, Mccurdy RD, Khokhar JY. Adolescent Substance Use and the Brain: Behavioral, Cognitive and Neuroimaging Correlates. Front Hum Neurosci 2020;14. 10.3389/fnhum.2020.00298.

[13] Everitt BJ, Robbins TW. Neural systems of reinforcement for drug addiction: from actions to habits to compulsion. Nat Neurosci 2005;8:1481–9. 10.1038/nn1579.

[14] Morein-Zamir S, Robbins TW. Fronto-striatal circuits in response-inhibition: Relevance to addiction. Brain Res 2015;1628:117–29. 10.1016/j.brainres.2014.09.012.

[15] Kim-Spoon J, Kahn RE, Lauharatanahirun N, Deater-Deckard K, Bickel WK, Chiu PH, et al. Executive Functioning and Substance Use in Adolescence: Neurobiological and Behavioral Perspectives. Neuropsychologia 2017;100:79–92. 10.1016/j.neuropsychologia.2017.04.020.

[16] Chinchella N, Hipólito I. Substance addiction: cure or care? Phenomenol Cogn Sci 2023. 10.1007/s11097-023-09885-3.

[17] Heilig M, MacKillop J, Martinez D, Rehm J, Leggio L, Vanderschuren LJMJ. Addiction as a brain disease revised: why it still matters, and the need for consilience. Neuropsychopharmacol 2021 4610 2021;46:1715–23. 10.1038/s41386-020-00950-y.

[18] Volkow ND, Koob G. Brain disease model of addiction: why is it so controversial? Lancet Psychiatry 2015;2:677–9. 10.1016/S2215-0366(15)00236-9.

[19] Wise RA. Addiction Becomes a Brain Disease. Neuron 2000;26:27–33. 10.1016/S0896-6273(00)81134-4.

[20] Lui CK, Witbrodt J, Li L, Tam CC, Williams E, Guo Z, et al. Associations between early childhood adversity and behavioral, substance use, and academic outcomes in childhood through adolescence in a U.S. longitudinal cohort. Drug Alcohol Depend 2023;244:109795. 10.1016/j.drugalcdep.2023.109795.

[21] Hoffmann JP, Jones MS. Cumulative Stressors and Adolescent Substance Use: A Review of 21st-Century Literature. Trauma Violence Abuse 2022;23:891–905. 10.1177/1524838020979674.

[22] Ramírez M, Ugedo A, Fañanás L, Cano-Escalera G, Saiz PAA, Zorrilla I, et al. Scoping review of biological and psychosocial pathways that lead from childhood adversity to early-onset substance use. Front Psychiatry 2025;16. 10.3389/fpsyt.2025.1612494.

[23] Watts LL, Hamza EA, Bedewy DA, Moustafa AA. A meta-analysis study on peer influence and adolescent substance use. Curr Psychol 2024;43:3866–81. 10.1007/s12144-023-04944-z.

[24] Coronado C, Wade NE, Aguinaldo LD, Hernandez Mejia M, Jacobus J. Neurocognitive Correlates of Adolescent Cannabis Use: an Overview of Neural Activation Patterns in Task-Based Functional MRI Studies. J Pediatr Neuropsychol 2020;6:1–13. 10.1007/s40817-020-00076-5.

[25] Cservenka A, Brumback T. The Burden of Binge and Heavy Drinking on the Brain: Effects on Adolescent and Young Adult Neural Structure and Function. Front Psychol 2017;8. 10.3389/fpsyg.2017.01111.

[26] Sullivan RM, Wallace AL, Shankula CA, Celhay O, Ziemer LR, Smith CJ, et al. Substance use patterns from late childhood to mid-adolescence: Updates on the Adolescent Brain Cognitive Development Study. Drug Alcohol Depend Rep 2026;20:100463. 10.1016/j.dadr.2026.100463.

[27] ABCD Consortium. ABCD Data Documentation (all versions) 2026. 10.5281/zenodo.15800724.

[28] Casey BJ, Cannonier T, Conley MI, Cohen AO, Barch DM, Heitzeg MM, et al. The Adolescent Brain Cognitive Development (ABCD) study: Imaging acquisition across 21 sites. Dev Cogn Neurosci 2018;32:43–54. 10.1016/j.dcn.2018.03.001.

[29] Hagler DJ, Hatton SN, Cornejo MD, Makowski C, Fair DA, Dick AS, et al. Image processing and analysis methods for the Adolescent Brain Cognitive Development Study. NeuroImage 2019;202:116091. 10.1016/j.neuroimage.2019.116091.

[30] Knutson B, Adams CM, Fong GW, Hommer D. Anticipation of Increasing Monetary Reward Selectively Recruits Nucleus Accumbens. J Neurosci 2001;21:RC159– RC159. 10.1523/JNEUROSCI.21-16-j0002.2001.

[31] Yau W-YW, Zubieta J-K, Weiland BJ, Samudra PG, Zucker RA, Heitzeg MM. Nucleus Accumbens Response to Incentive Stimuli Anticipation in Children of Alcoholics: Relationships with Precursive Behavioral Risk and Lifetime Alcohol Use. J Neurosci 2012;32:2544–51. 10.1523/JNEUROSCI.1390-11.2012.

[32] Balodis IM, Potenza MN. Anticipatory Reward Processing in Addicted Populations: A Focus on the Monetary Incentive Delay Task. Biol Psychiatry 2015;77:434–44. 10.1016/j.biopsych.2014.08.020.

[33] Beck A, Schlagenhauf F, Wüstenberg T, Hein J, Kienast T, Kahnt T, et al. Ventral Striatal Activation During Reward Anticipation Correlates with Impulsivity in Alcoholics. Biol Psychiatry 2009;66:734–42. 10.1016/j.biopsych.2009.04.035.

[34] Wrase J, Schlagenhauf F, Kienast T, Wüstenberg T, Bermpohl F, Kahnt T, et al. Dysfunction of reward processing correlates with alcohol craving in detoxified alcoholics. NeuroImage 2007;35:787–94. 10.1016/j.neuroimage.2006.11.043.

[35] Hart H, Radua J, Nakao T, Mataix-Cols D, Rubia K. Meta-analysis of Functional Magnetic Resonance Imaging Studies of Inhibition and Attention in Attention-deficit/Hyperactivity Disorder: Exploring Task-Specific, Stimulant Medication, and Age Effects. JAMA Psychiatry 2013;70:185–98. 10.1001/jamapsychiatry.2013.277.

[36] Logan GD, Van Zandt T, Verbruggen F, Wagenmakers E-J. On the ability to inhibit thought and action: General and special theories of an act of control. Psychol Rev 2014;121:66–95. 10.1037/a0035230.

[37] Whelan R, Conrod PJ, Poline J-B, Lourdusamy A, Banaschewski T, Barker GJ, et al. Adolescent impulsivity phenotypes characterized by distinct brain networks. Nat Neurosci 2012;15:920–5. 10.1038/nn.3092.

[38] Boen R, Raud L, Huster RJ. Inhibitory Control and the Structural Parcelation of the Right Inferior Frontal Gyrus. Front Hum Neurosci 2022;16. 10.3389/fnhum.2022.787079.

[39] Breslin FJ, Ratliff EL, Cohen ZP, Croff JM, Kerr KL. Measuring adversity in the ABCD® Study: systematic review and recommendations for best practices. BMC Med Res Methodol 2025;25:77. 10.1186/s12874-025-02521-5.

[40] Johnston LD, O’Malley PM, Bachman JG, Schulenberg JE, Miech RA. Monitoring the Future National Survey Results on Drug Use, 1975-2013. Volume 1, Secondary School Students. Institute for Social Research; 2014.

[41] Johnston LD, Others A. Illicit Drug Use, Smoking, and Drinking by America’s High School Students, College Students, and Young Adults 1975–1987. Superintendent of Documents, U; 1988.

[42] Lisdahl KM, Sher KJ, Conway KP, Gonzalez R, Feldstein Ewing SW, Nixon SJ, et al. Adolescent brain cognitive development (ABCD) study: Overview of substance use assessment methods. Dev Cogn Neurosci 2018;32:80–96. 10.1016/j.dcn.2018.02.007.

[43] PhenX Toolkit n.d. https://www.phenxtoolkit.org/ (accessed June 18, 2026).

[44] Posit team. RStudio: Integrated Development Environment for R 2025.

[45] R Core Team. R: A Language and Environment for Statistical Computing 2024.

[46] Therneau TM, Thomas Lumley (original S.->R port and R. maintainer until 2009), Elizabeth A, Cynthia C. survival: Survival Analysis 2026.

[47] Wickham H. ggplot2: Elegant Graphics for Data Analysis 2016.

[48] Wickham H, François R, Henry L, Müller K. dplyr: A Grammar of Data Manipulation 2018.

[49] Austin PC, Giardiello D. The Impact of Violation of the Proportional Hazards Assumption on the Calibration of the Cox Proportional Hazards Model. Stat Med 2025;44:e70161. 10.1002/sim.70161.

[50] Hughes ME, Johnston PJ, Fulham WR, Budd TW, Michie PT. Stop-signal task difficulty and the right inferior frontal gyrus. Behav Brain Res 2013;256:205–13. 10.1016/j.bbr.2013.08.026.

[51] Chaarani B, Hahn S, Allgaier N, Adise S, Owens MM, Juliano AC, et al. Baseline brain function in the preadolescents of the ABCD Study. Nat Neurosci 2021;24:1176–86. 10.1038/s41593-021-00867-9.

[52] White AM. Gender Differences in the Epidemiology of Alcohol Use and Related Harms in the United States. Alcohol Res Curr Rev 2020;40:01. 10.35946/arcr.v40.2.01.

[53] Chapman C, Slade T, Swift W, Keyes K, Tonks Z, Teesson M. Evidence for Sex Convergence in Prevalence of Cannabis Use: A Systematic Review and Meta-Regression. J Stud Alcohol Drugs 2017;78:344–52. 10.15288/jsad.2017.78.344.

[54] Cuttler C, Mischley LK, Sexton M. Sex Differences in Cannabis Use and Effects: A Cross-Sectional Survey of Cannabis Users. Cannabis Cannabinoid Res 2016;1:166–75. 10.1089/can.2016.0010.

[55] Miech RA, Patrick ME, Jager JO, Jang JB. Monitoring the future national survey results on drug use, 1975–2025: Overview and detailed results for secondary school students. Institute for Social Research, University of Michigan; 2026.

[56] Health Canada. Canadian Tobacco Alcohol and Drugs (CTADS): 2015 summary 2017. https://www.canada.ca/en/health-canada/services/canadian-alcohol-drugs-survey/2015-summary.html (accessed June 26, 2026).

[57] Health Canada. Alcohol and Drug Use among Students in Canada, 2023–24 2025. https://www.canada.ca/en/health-canada/services/canadian-student-tobacco-alcohol-drugs-survey/2023-2024-key-findings.html (accessed June 26, 2026).

[58] Hanson MD, Chen E. Socioeconomic Status and Substance Use Behaviors in Adolescents: The Role of Family Resources versus Family Social Status. J Health Psychol 2007;12:32–5. 10.1177/1359105306069073.

[59] Humensky JL. Are adolescents with high socioeconomic status more likely to engage in alcohol and illicit drug use in early adulthood? Subst Abuse Treat Prev Policy 2010;5:19. 10.1186/1747-597X-5-19.

[60] Jang JB, Patrick ME, Keyes KM, Hamilton AD, Schulenberg JE. Frequent Binge Drinking Among US Adolescents, 1991 to 2015. Pediatrics 2017;139:e20164023. 10.1542/peds.2016-4023.

[61] Martz ME, Heitzeg MM, Lisdahl KM, Cloak CC, Ewing SWF, Gonzalez R, et al. Individual-, peer-, and parent-level substance use-related factors among 9- and 10-year-olds from the ABCD Study: Prevalence rates and sociodemographic differences. Drug Alcohol Depend Rep 2022;3:100037. 10.1016/j.dadr.2022.100037.

[62] Tolstrup JS, Kruckow S, Becker U, Andersen O, Sawyer SM, Katikireddi SV, et al. Socioeconomic inequalities in alcohol-related harm in adolescents: a prospective cohort study of 68,299 Danish 15–19-year-olds. eClinicalMedicine 2023;62. 10.1016/j.eclinm.2023.102129.

[63] Bray JH, Adams GJ, Getz JG, McQueen A. Individuation, peers, and adolescent alcohol use: A latent growth analysis. J Consult Clin Psychol 2003;71:553–64. 10.1037/0022-006X.71.3.553.

[64] Curran PJ, Stice E, Chassin L. The relation between adolescent alcohol use and peer alcohol use: A longitudinal random coefficients model. J Consult Clin Psychol 1997;65:130–40. 10.1037/0022-006X.65.1.130.

[65] Henneberger AK, Mushonga DR, Preston AM. Peer Influence and Adolescent Substance Use: A Systematic Review of Dynamic Social Network Research. Adolesc Res Rev 2021;6:57–73. 10.1007/s40894-019-00130-0.

[66] McDonough MH, Jose PE, Stuart J. Bi-directional Effects of Peer Relationships and Adolescent Substance Use: A Longitudinal Study. J Youth Adolesc 2016;45:1652– 63. 10.1007/s10964-015-0355-4.

[67] Simons-Morton B, Chen RS. Over time relationships between early adolescent and peer substance use. Addict Behav 2006;31:1211–23. 10.1016/j.addbeh.2005.09.006.

[68] Smit K, Voogt C, Hiemstra M, Kleinjan M, Otten R, Kuntsche E. Development of alcohol expectancies and early alcohol use in children and adolescents: A systematic review. Clin Psychol Rev 2018;60:136–46. 10.1016/j.cpr.2018.02.002.

[69] Bahr SJ, Hoffmann JP, Yang X. Parental and Peer Influences on the Risk of Adolescent Drug Use. J Prim Prev 2005;26:529–51. 10.1007/s10935-005-0014-8.

[70] Farley JP, Kim-Spoon J. Longitudinal Associations among Impulsivity, Friend Substance Use, and Adolescent Substance Use. J Addict Res Ther 2015;6:1000220. 10.4172/2155-6105.1000220.

[71] Kim-Spoon J, Deater-Deckard K, Lauharatanahirun N, Farley JP, Chiu PH, Bickel WK, et al. Neural Interaction Between Risk Sensitivity and Cognitive Control Predicting Health Risk Behaviors Among Late Adolescents. J Res Adolesc Off J Soc Res Adolesc 2017;27:674–82. 10.1111/jora.12295.

[72] Villafuerte S, Heitzeg MM, Foley S, Wendy Yau W-Y, Majczenko K, Zubieta J-K, et al. Impulsiveness and insula activation during reward anticipation are associated with genetic variants in GABRA2 in a family sample enriched for alcoholism. Mol Psychiatry 2012;17:511–9. 10.1038/mp.2011.33.

[73] Feldstein Ewing SW, Dash GF, Thompson WK, Reuter C, Diaz VG, Anokhin A, et al. Measuring retention within the adolescent brain cognitive development (ABCD)SM study. Dev Cogn Neurosci 2022;54:101081. 10.1016/j.dcn.2022.101081.

[74] Saragosa-Harris NM, Chaku N, MacSweeney N, Guazzelli Williamson V, Scheuplein M, Feola B, et al. A practical guide for researchers and reviewers using the ABCD Study and other large longitudinal datasets. Dev Cogn Neurosci 2022;55:101115. 10.1016/j.dcn.2022.101115.

